# Self-supervised neural network improves tri-exponential intravoxel incoherent motion model fitting in non-alcoholic fatty liver disease

**DOI:** 10.1101/2022.04.04.22273378

**Authors:** Marian A. Troelstra, Anne-Marieke van Dijk, Julia J. Witjes, Anne Linde Mak, Diona Zwirs, Jurgen H. Runge, Joanne Verheij, Ulrich H. Beuers, Max Nieuwdorp, Adriaan G. Holleboom, Aart J. Nederveen, Oliver J. Gurney-Champion

**Author notes:** Corresponding author Marian Amber Troelstra, Department of Radiology and Nuclear Medicine, Amsterdam UMC, location AMC, Meibergdreef 9, 1105AZ Amsterdam, The Netherlands.

## Abstract

Neural network generated intravoxel incoherent motion (IVIM) parameter maps are visually more detailed, less noisy and have a higher test-retest repeatability than conventional least-squares (LSQ) generated images. Recent literature suggests that tri-exponential models may fit liver IVIM data more accurately than conventional bi-exponential models, warranting investigation into the use of a neural network (NN) to estimate tri-exponential IVIM parameters. Here, we developed a tri-exponential IVIM unsupervised physics-informed deep neural network (IVIM_3_-NET), assessed its performance in non-alcoholic fatty liver disease (NAFLD) and compared outcomes with bi-exponential LSQ and NN fits and tri-exponential LSQ fits. Scanning was performed using a 3.0T free-breathing multi-slice diffusion-weighted single-shot echo-planar imaging sequence with 18 b-values. Images were analysed for visual quality, comparing the bi- and tri-exponential IVIM models for LSQ fits and NN fits using parameter-map signal-to-noise ratios (SNR) and adjusted R^2^. IVIM parameters were compared to histological fibrosis, disease activity and steatosis grades. Parameter map quality improved with bi- and tri-exponential NN approaches, with average parameter-map SNR increasing from 3.38 to 5.59 and 2.45 to 4.01 for bi- and tri-exponential LSQ and NN models respectively. In 33 out of 36 patients, the tri-exponential model exhibited higher adjusted R^2^ values than the bi-exponential model. Correlating IVIM data to liver histology showed that the bi- and tri-exponential NN outperformed both LSQ models for the majority of IVIM parameters (10 out of 15 significant correlations). Overall, our results support the use of a tri-exponential IVIM model in NAFLD. We show that the IVIM_3_-NET can be used to improve image quality compared to a tri-exponential LSQ fit and provides promising correlations with histopathology.

## Introduction

The prevalence and degree of obesity and type 2 diabetes mellitus are increasing worldwide, resulting in a concomitant increase in non-alcoholic fatty liver disease (NAFLD), which is now the most prevalent global liver disease^1^. NAFLD is characterized by an accumulation of lipids within hepatocytes, known as steatosis, which in turn can trigger an inflammatory response in the liver, leading to hepatocyte ballooning, lobular inflammation and ultimately into liver fibrosis^2^. NAFLD can lead to liver-related complications such as cirrhosis and hepatocellular carcinoma^1^, and is also thought to contribute to an increased risk of atherosclerotic cardiovascular disease^3,4^. Liver biopsy is currently the gold standard for diagnosing and staging NAFLD, assessing levels of steatosis, inflammation, ballooning and fibrosis from a small tissue sample. Several disadvantages of liver biopsy include patient discomfort, a small, yet severe risk of life-threatening intraperitoneal haemorrhage and risk of sampling error^5,6^. This has led to a growing interest in non-invasive techniques for detecting the presence and severity of NAFLD. MRI has been developed and researched as a diagnostic tool in NAFLD^7^. Various methods have been proposed, of which intravoxel incoherent motion (IVIM) imaging has shown potential as a biomarker for assessing and staging NAFLD, particularly for levels of fibrosis^8^.

IVIM assesses tissue diffusivity and perfusion reflected by random motion of water molecules in the intracellular and extracellular spaces as well as in the tissue (micro)circulation, respectively^9^. Data acquired using a range of diffusion weightings (b-values) are typically fitted using a bi-exponential model using the following formula:

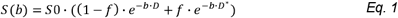

where diffusion (*D*), pseudo-diffusion (*D*\*) and perfusion fraction (*f*) parameters are estimated, with *S*0 equating the signal intensity for b=0 s/mm^2^. NAFLD has the potential to alter IVIM parameters in multiple ways. In the case of fibrosis, it can be theorized that the increased collagen depositions in the liver restrict the free movement of water molecules, leading to a reduction in the diffusion parameters, while increased hepatic resistance amongst others can lead to a reduction in hepatic perfusion. Li et al. show that while absolute values tend to vary between studies, typically a decrease in all three IVIM parameters is observed as fibrosis stage increases^8^. Hepatic inflammation could be thought to decrease perfusion parameters in particular due to the inflammatory response, in which cell infiltration and oedema restrict blood flow in the capillaries. Hepatocellular ballooning leads to increases in cell size and therefore in theory could restrict both diffusion and perfusion parameters. While the literature on IVIM for assessing inflammation or ballooning grades is less readily available, the existing literature describes a negative correlation between perfusion fraction and both inflammation and ballooning^10^.

Recent literature suggests that the bi-exponential IVIM model may be insufficient for fitting the liver’s complex structure. The marked signal decay in the IVIM signal intensity plots seen at low b-values has given rise to the hypothesis that a tri-exponential model will provide a more accurate representation of the data and improve fit accuracy^11^. Herein, an extra exponent is added to improve this fitting of rapid signal decay at very low b-values:

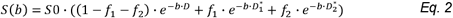

where diffusion (*D*), slow 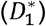 and fast pseudo-diffusion 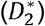, and slow (*f*_1_) and fast perfusion fraction (*f*_2_) are estimated. Available studies advocating the use of a tri-exponential model show a higher goodness of fit when compared to a bi-exponential model^12–14^, seen by an improved Akaike information criterion amongst others. The tri-exponential model with an extra fast diffusion component, however, has only been implemented in a limited number of studies and thus far only in healthy volunteers. Its added value for disease classification, such as in patients with NAFLD, yet needs to be investigated.

Although tri-exponential fitting more accurately describes the data, fitting of this complex model is challenging and current least-squares (LSQ) methods typically lead to noisy parameter maps, particularly in the case of pseudo-diffusion. As a result, tri-exponential fits are often done using averaged data from a region of interest (ROI), or even averaged data from multiple subjects, instead of voxel-wise fitting, reducing clinical diagnostic applicability.

To improve parameter map accuracy, the use of deep neural networks to model the bi-exponential IVIM fit has been proposed^15–19^. In a previous study, a physics-informed unsupervised approach that could train directly to in-vivo MRI data (IVIM-NET)^15,20^ was implemented. IVIM-NET provided parameter maps that were less noisy, visually more detailed and had a higher test-retest repeatability. However, there is not yet a tri-exponential equivalent to the neural network. This approach could substantially improve the voxel-wise fitting of tri-exponential IVIM, enabling parameter maps with clinically acceptable quality and aid wider spread implementation.

Therefore, in this work, we aim to investigate the value of a deep neural network to fit a tri-exponential model to IVIM data in NAFLD. We split up the research into two sub-questions: firstly, does the parameter map quality improve when introducing neural networks for fitting the bi- and tri-exponential models? Secondly, is there an added value of tri-exponential fitting in the non-invasive grading of NAFLD disease severity? To address these questions, we have developed a deep neural network to fit a tri-exponential model to IVIM data (IVIM_3_-NET). We have tested the tri-exponential versus bi-exponential model in patients with various stages of NAFLD, and compared the use of IVIM-NET and IVIM_3_-NET to conventional bi- and tri-exponential LSQ fits.

## Experimental

### Design

Patient data were collected as part of the ongoing Amsterdam NAFLD-NASH cohort (ANCHOR) study as described in previous work^21^. In this previous clinical work, the correlations between liver histopathology and several ultrasonographic and MRI parameters, including the bi-exponential IVIM-model, were studied. In our current technical work, we use the data to study the tri-exponential model and the performance of NNs. All patients underwent an MRI scan of the liver and ultrasound-guided liver biopsy, amongst other diagnostic tests. The study was conducted at Amsterdam University Medical Centre, in compliance with the principles in the declaration of Helsinki and according to Good Clinical Practice guidelines. The protocol was reviewed and approved by the institutional review board of the AMC and was registered in the Dutch Trial Register (registration number NTR7191). All participants provided written informed consent before study activities.

### Participants

36 individuals with known hepatic steatosis (confirmed on ultrasound), a BMI > 25kg/m^2^ and elevated ASAT and/or ALAT levels were included. The main exclusion criteria were contraindications for undergoing MRI, any condition or risk factors potentially leading to liver disease besides NAFLD (excessive alcohol use, drug-use, hepatitis, etc.), conditions or risk factors leading to bleeding disorders and decompensated liver cirrhosis or hepatocellular carcinomas. Participants were required to fast for a minimum of 4 hours before diagnostic tests.

### MRI acquisition

MRI scanning was performed using a 3.0T MRI scanner (Ingenia; Philips, Best, The Netherlands) with a 16-channel phased-array anterior coil and 10-channel phase-arrayed posterior coil. IVIM data were acquired using a free-breathing multi-slice diffusion-weighted single-shot echo-planar imaging sequence. Eighteen unique b*-*values were acquired: 0, 1, 2, 5, 10, 20, 30, 40, 50, 75, 100, 150, 200, 300, 400, 500, 600, and 700 s/mm^2^ in three orthogonal directions. The following acquisition parameters were used: repetition time 7000 ms, echo time 45.5 ms, bandwidth 20.8 Hz/pixel in phase encoding direction, field-of-view 450×295 mm^2^ with a 3.0×3.0 mm^2^ acquisition resolution, 27 slices with a 6.0 mm slice thickness and 1.0 mm slice gap, parallel imaging factor (SENSE) 1.3, partial averaging factor 0.6, spectral attenuated inversion recovery (SPAIR) fat suppression and three saturation slabs to suppress signal from the anterior abdominal wall. The total acquisition time was 8.1 minutes.

### MRI analysis

Images were analysed by a single observer (M.T.) with four years of experience in hepatic MRI. The entire liver was manually segmented using reconstructed b-value = 0 images, avoiding liver edges and any areas or slices containing (motion) artefacts (ROI_liver_). Four different models were used to fit the ROI_liver_, all implemented in Python (version 3.6.12). These consisted of our newly developed IVIM_3_-NET tri-exponential fit, the IVIM-NET bi-exponential fit, and the LSQ bi- and tri-exponential fits. Data were normalized to the S(b=0 s/mm^2^) for all fitting methods. For the deep learning approaches, we used PyTorch (1.8.1) whereas the least-squares fitting was done with scipy’s (1.5.2) optimize.curve_fit function. All fitting methods can be found on https://github.com/oliverchampion/IVIMNET. From these segmented images a small ROI (ROI_SNR_) in the right liver lobe in homogenous liver tissue.

#### Bi- and tri-exponential LSQ fits

The LSQ models were fit voxel-wise, solving the bi- or tri-exponential model for all segmented voxels. For the bi-exponential LSQ fitting, Eq. 1 was fitted directly, with fit constraints 3×10^−4^<*D*<5×10^−3^ mm^2^/s, 0<*f* <0.7, 5×10^−3^<*D*\*<3×10^−1^ mm^2^/s and 0.5<*S*0 <2.5 mm^2^/s. For the tri-exponential LSQ fitting, Eq. 2 was modified and we fitted:

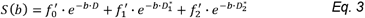

with 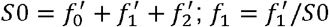; and 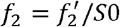.

Constraints were implemented ensuring 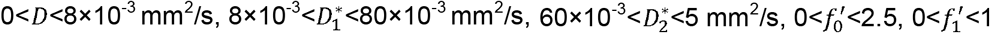 and 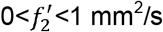. The *f*_1_ and *f*_2_ of the tri-exponential model were also summed to produce a *f*_1+2_ parameter map for direct comparison with the perfusion fraction *f* of the bi-exponential model.

To determine goodness of fit, the bi- and tri-exponential LSQ models were also separately fit in a ROI-wise manner using the ROI_liver_. Here the average value per individual and per b-value was fit using the same constraints as described above, increasing the signal-to-noise ratio (SNR). Fitting was done in a parallel routine, using 8 cores from an Intel(R) Xeon(R) Gold 6132 CPU @ 2.60GHz and fitting times were recorded.

#### IVIM-NET and IVIM_3_-NET fits

IVIM-NET bi and tri-exponential fits were also performed voxel-wise. The IVIM-NET bi-exponential fit was performed following previously published work, using a fully-connected network per IVIM parameter (*S*0, *D, D*\* and *f*) and a physics-informed loss function to minimize the root-mean-square error between input voxel signals and predicted signal decay. The hyperparameters were kept identical to the optimized setting previously published^16^, with a dropout of 0.1, batch normalization, 2 hidden layers, Adam optimizer and a learning rate of 3×10^−5^.

The tri-exponential IVIM_3_-NET was developed based on the IVIM-NET, adapting the physics-informed loss function to a tri-exponential model (Eq. 3, Figure 1), instead of the previously reported bi-exponential model. As the tri-exponential model is more challenging to fit than the bi-exponential model, we made some additional adaptations to the network. We introduced a semi-parallel training approach, which was a variation on the parallel training from IVIM-NET. Instead of having each parameter estimated by a separate network, however, each pair of decay constants and signal fractions (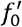 with 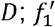 and 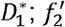 and 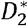) were predicted by a network. We used 4 hidden layers (instead of 2). We also started at a higher learning rate of 1×10^−4^ and introduced a scheduler that decreased the learning rate by a factor of 5 each time the network did not improve performance on the validation data for 10 consecutive iterations. Early stopping was then adapted such that once the performance did not decrease in the initial 10 iterations of a new learning rate, training was stopped.

**Figure 1.**
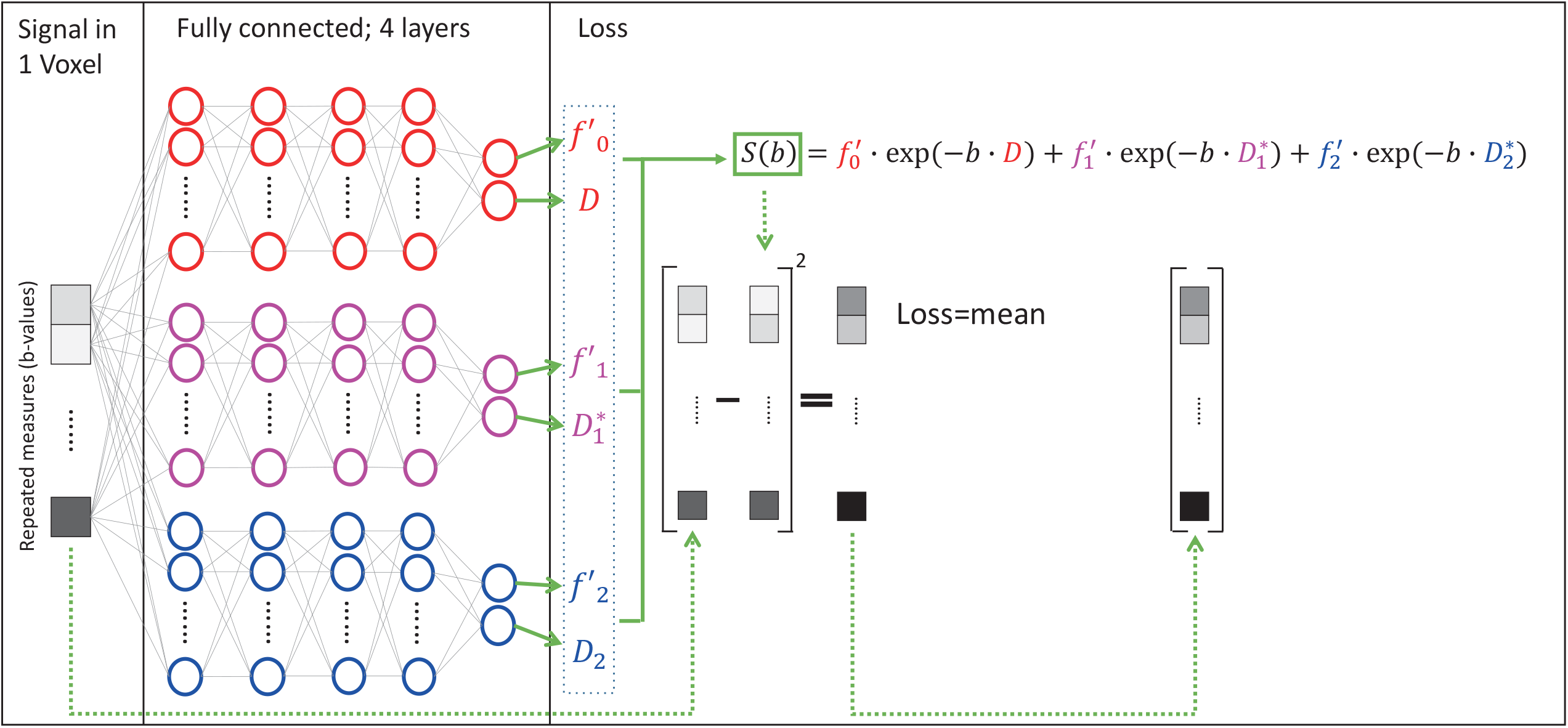
Deep neural network for fitting of a tri-exponential model to IVIM data (IVIM_3_-NET). The IVIM_3_-NET fits a tri-exponential model to the data, solving the equation per voxel.

Both networks were fitted to the same in vivo liver data, from all patients, resulting in 3,304,437 voxels. Data were split into 90% for training and 10% for testing. Per epoch, only a subset of 500 randomly selected batches (<2% of all data) were shown, while all testing data was tested between epochs. Training and inference were done on a Tesla P100 GPU and times were recorded (averaged over 10 repeated training/inference).

### Liver biopsy

Percutaneous ultrasound-guided liver biopsies were assessed by an experienced liver pathologist (J.V.). Histology specimens were scored according to the steatosis, activity and fibrosis score^22^. Steatosis levels were scored accordingly for grades S0-S3. NAFLD disease activity comprised the combined sum of inflammation grade (0-2) and ballooning grade (0-2), for a combined score ranging from grade A0-A4. Fibrosis was scored according to the NASH Clinical Research Network for severity ranging from grades F0-F4^23^.

### Statistical analysis

The statistical analysis was performed using R version 3.6.3^24^.

The SNR of the parameter maps in homogeneous tissue was used to determine whether the NN fitting approach was less noisy than the LSQ approach. A separate ROI (ROI_SNR_) was selected in the right liver lobe in homogenous liver tissue, avoiding liver edges, (motion) artefacts and visible vessels. For each individual, the SNR was defined as the ratio of the mean parameter value within ROI_SNR_ and the standard deviation from ROI_SNR_. Differences between means and SNRs of the bi- and tri-exponential models were assessed using Wilcoxon signed-rank tests. A p-value < 0.05 was considered statistically significant.

To determine whether the bi- or tri-exponential model more accurately described the data, we determined the adjusted R^2^ of the least-squares fit of the models. As the bi- and tri-exponential models have a different degree of freedom, we used the adjusted R^2^ which corrects for the extra degrees of freedom. As the noise was dominating the voxel-wise adjusted R^2^, we opted for an ROI-wise fit from the ROI_liver_, finding the adjusted R^2^ for each individual.

Comparison of histology and the average IVIM parameter values from the ROI_liver_ were performed using Spearman’s rank correlation for all four models. IVIM_3_-NET parameters were further assessed using Kruskall-Wallis tests for trends, and where appropriate followed by Dunn’s post-hoc analysis with p-value adjustment according to the Holm–Bonferroni method. A p-value < 0.05 was considered statistically significant.

## Results

### Patient characteristics

The first thirty-six individuals (22 male, 14 female) included in the ANCHOR study were analysed in this work. Liver fibrosis was found on biopsy in all but two patients (F0). Seven patients were classified as F1, nineteen as F2, seven as F3 and one as F4. All patients showed a steatosis level ≥1 on biopsy, of which 20 patients had S1, twelve S2 and four S3. Three patients had a disease activity grade of A0, twelve A1, fifteen A2, five A3 and one A4.

### Image quality

Visual assessment of IVIM parameter maps revealed that both the neural network methods showed an increased image quality and improved fit accuracy compared to the LSQ methods (Figure 2). The tri-exponential fit successfully provided parameter maps for all IVIM parameters, with a distinct difference between the 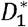 and 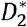 parameters as well as between *f*_1_ and *f*_2_. Peripheral vessels and liver tissue (e.g. capillaries) showed high 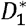 and *f*_1_ signals, while 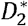 and *f*_2_ signals were high in larger vessels. In cases with a lower quality of acquired data, the IVIM_3_-NET was able to provide smoother parameter maps with less extreme outliers than the LSQ fit (Figure 3). Figure 4 displays IVIM_3_-NET parameter maps for three patients with increasing fibrosis grades. While all parameters except 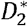 show a decrease in signal with increasing fibrosis grade, this effect is most noticeable for the perfusion parameter maps (*f*_1_, *f*_2_ and *f*_1+2_).

**Figure 2.**
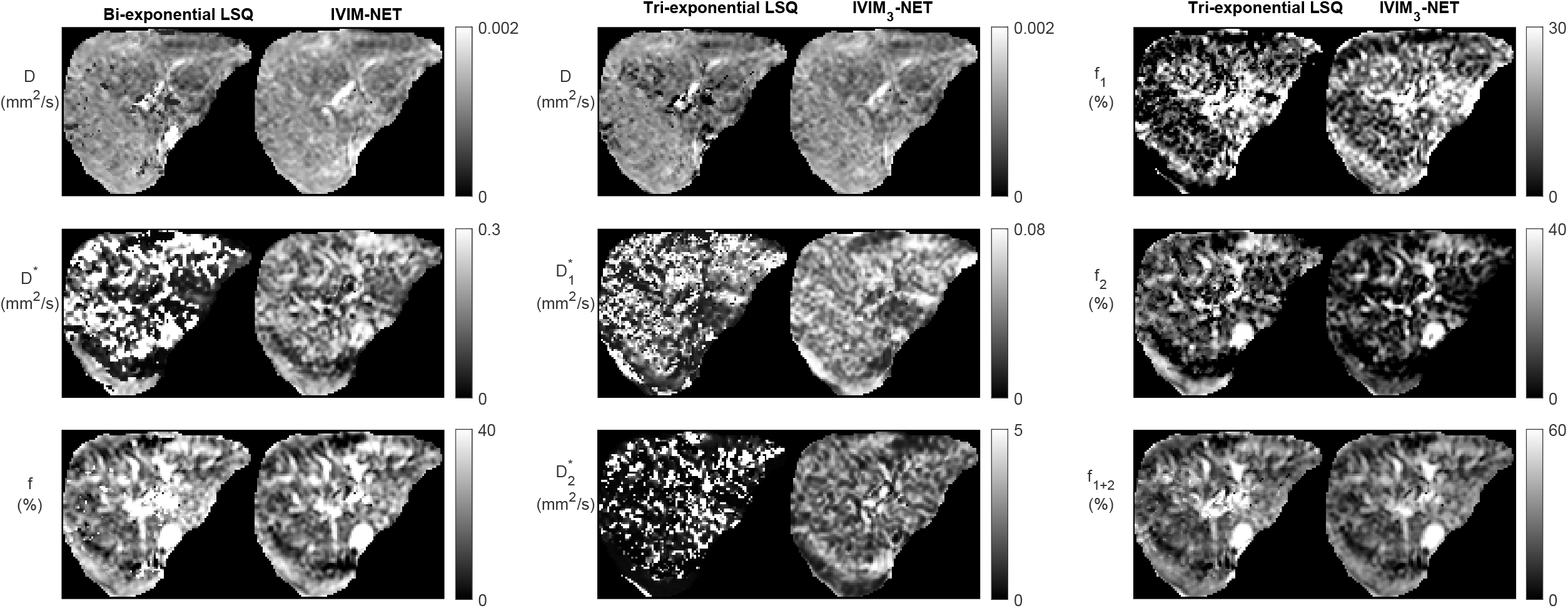
Example datasets from a single participant depicting the bi-exponential least-squares fit versus the bi-exponential neural network IVIM-NET fit on the left and tri-exponential least-squares fit versus tri-exponential neural network IVIM_3_-NET fit. Both neural network methods show less noisy parameter maps compared to the least-squares fits.

**Figure 3.**
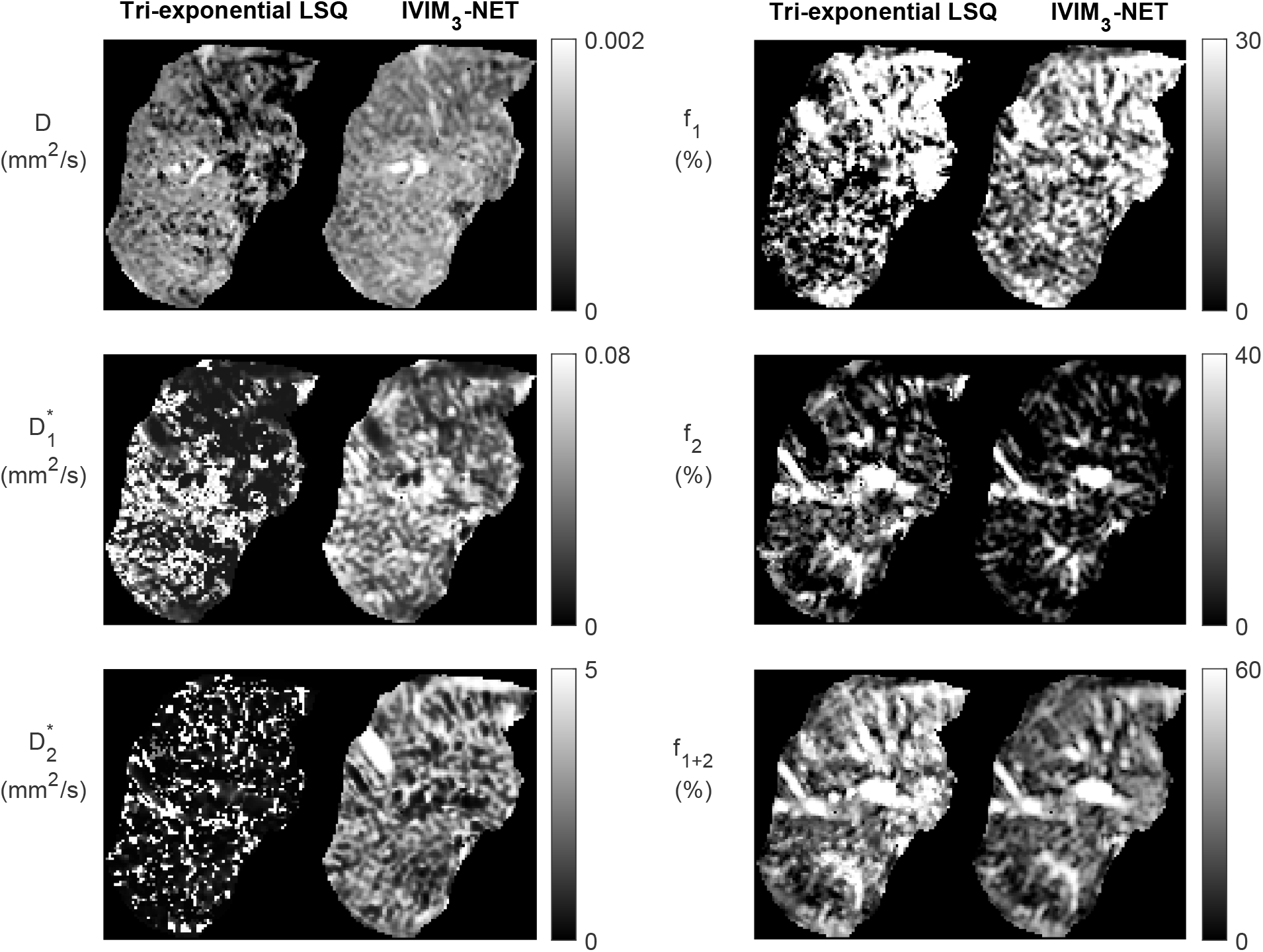
Example of a noisy dataset from a single participant with lower quality of image acquisition, fit with the tri-exponential least-squares fit and the tri-exponential neural network IVIM_3_-NET. The IVIM_3_-NET provides less noisy parameter maps.

**Figure 4.**
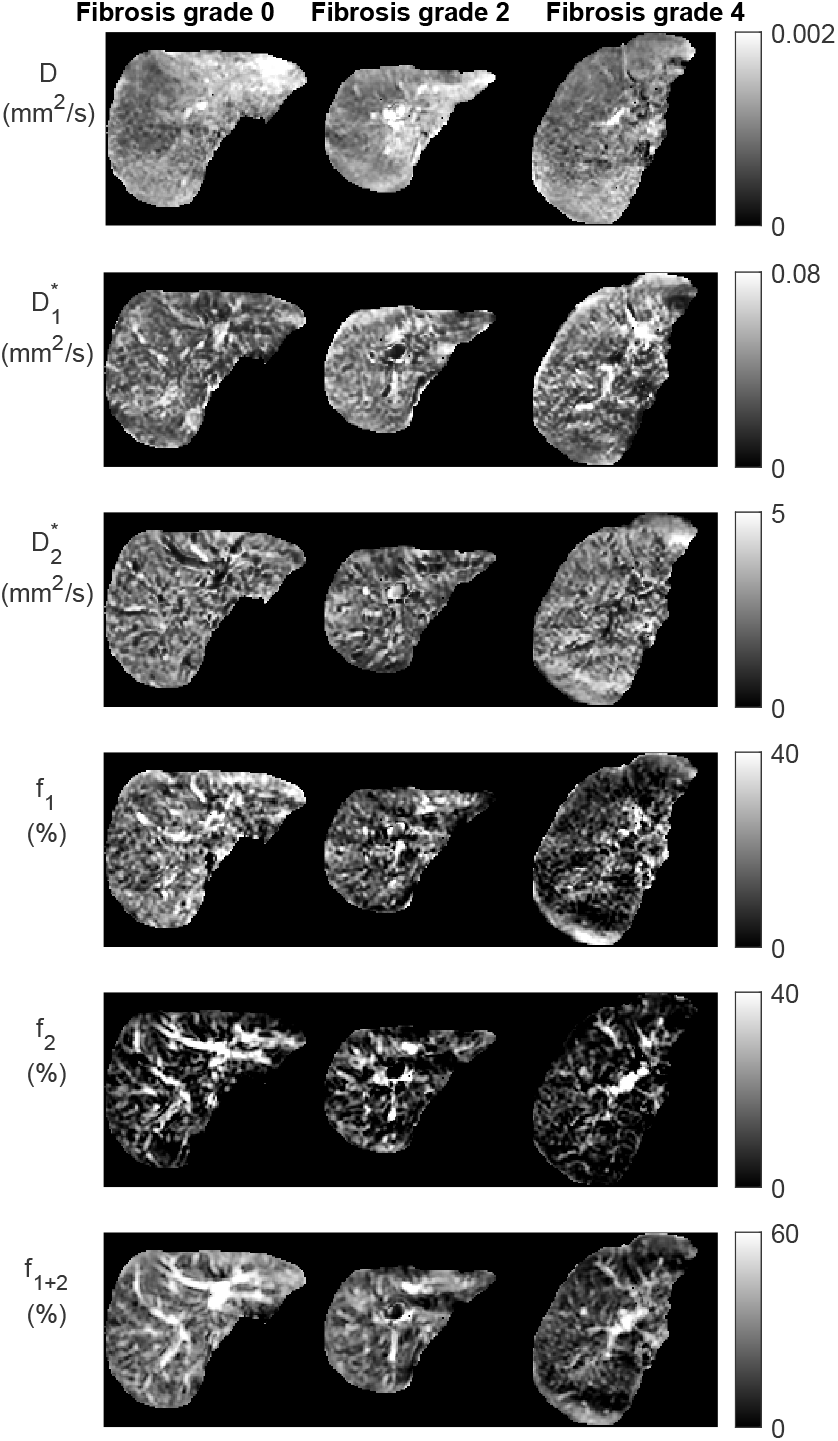
Datasets from the IVIM_3_-NET tri-exponential neural network of three patients with increasing levels of fibrosis. Visually a decrease of signal intensity with increasing fibrosis grade is most noticeable for the perfusion fractions (*f*_l_, *f*_2_ and *f*_l+2_).

One patient showed two incidental findings in the liver (Figure 5). Compared to the tri-exponential LSQ fit, the IVIM_3_-NET provided a more distinct delineation of the abnormalities. Interestingly, the two findings displayed different behaviour. The large lesion was most clearly visible on the IVIM_3_-NET pseudo-diffusion maps, with a low 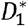 signal and a high 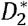 signal. On the other hand, the small lesion was clearer on the perfusion maps, with a low *f*_1_ signal in and a high *f*_2_ signal.

**Figure 5.**
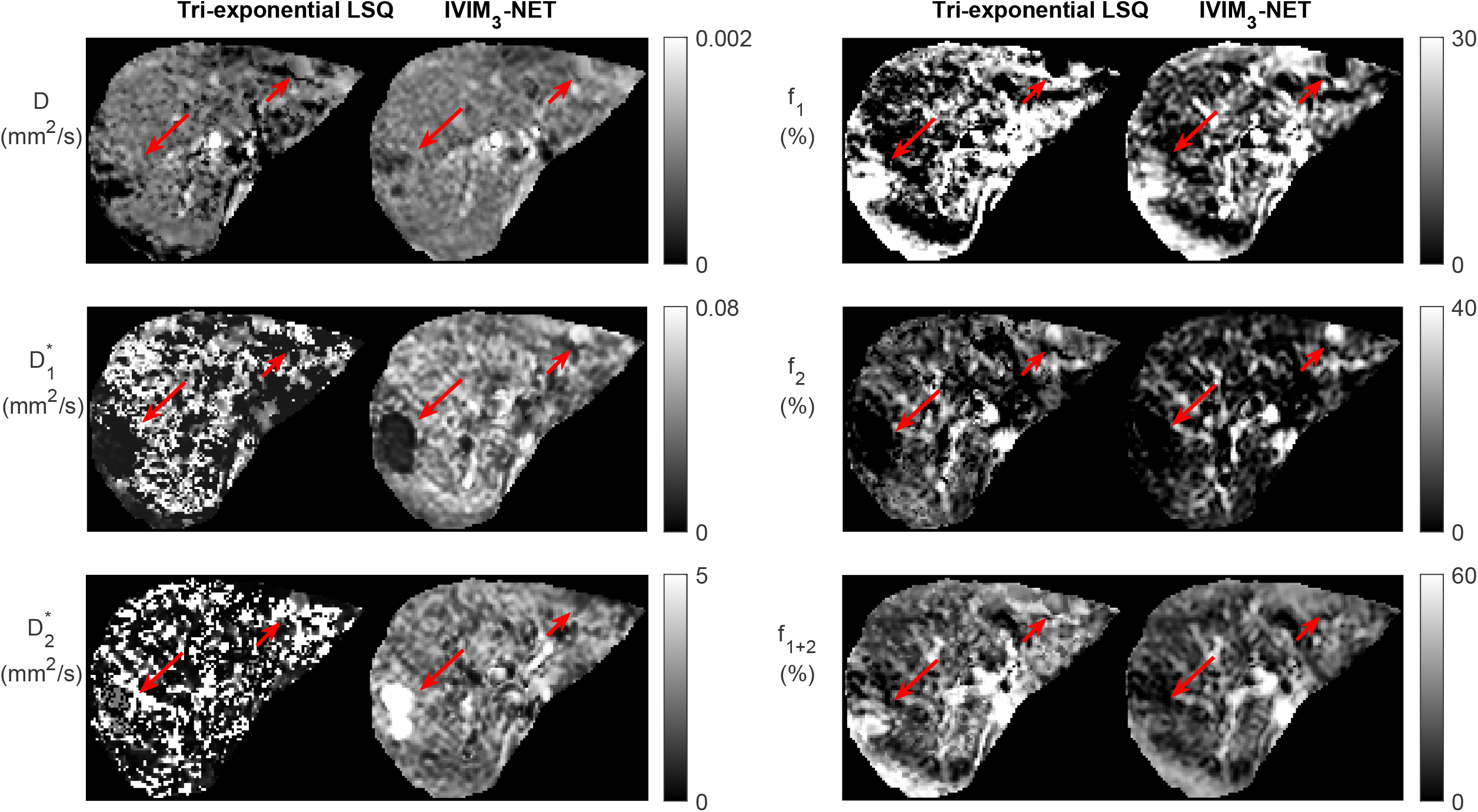
Dataset from individual with two incidental findings, as seen on the tri-exponential least-squares fit and the IVIM_3_-NET tri-exponential neural network fit and indicated by the red arrows. IVIM_3_-NET in particular shows a clear delineation of the lesions.

### Comparison of neural network versus least-squares model

The neural network approaches resulted in substantially less noisy, and hence more precise, parameter maps. The average spread in IVIM values from the ROI_SNR_ in homogenous liver tissue showed significantly higher SNRs in the parameter maps for both neural networks fits compared to the LSQ fits for all parameters except the tri-exponential *f*_2_ (Table 1 and Supplementary Information Figure S1). The average SNR over all parameters was 3.38 versus 5.59 for the bi-exponential LSQ and NN fit respectively and 2.45 versus 4.01 for the tri-exponential LSQ and NN fit respectively.

**Table 1.** Mean parameter values from the entire liver excluding large vessels (ROI_liver_) and mean signal-to-noise ratio (SNR) from the small region-of-interest in homogenous liver tissue (ROI_SNR_) for all four IVIM models per IVIM parameter. The bi-exponential neural network (NN) model had significantly higher means for the D and D* parameters, while the f was higher for bi-exponential least-squares (LSQ) model. The SNR was higher for all bi-exponential NN parameters. The tri-exponential NN model had a higher mean values for the 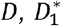 and 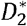 parameters; the tri-exponential LSQ model had higher means for the f_1_, f_2_ and f_l+2_ parameters. SNR of the tri-exponential NN model was significantly higher than the tri-exponential LSQ model for all parameters except f_2_ (LSQ higher than NN) and f_l+2_ (no significant difference between NN and LSQ). *: significantly higher SNR than other bi- or tri-exponential model with 0.01 < p-value < 0.05. **: significantly higher SNR than other bi- or tri-exponential model with 0.001 < p-value < 0.01. ***: significantly higher SNR than other bi- or tri-exponential model with p-value < 0.001. LSQ: least-squares, NN: neural network, SNR: signal-to-noise ratio.

|  | Bi-exponential LSQ |  | Bi-exponential NN |  | Tri-exponential LSQ |  | Tri-exponential NN |  |
| --- | --- | --- | --- | --- | --- | --- | --- | --- |
|  | Mean | SNR | Mean | SNR | Mean | SNR | Mean | SNR |
| $D$ ($10^{-3}$ mm<sup>2</sup>/s) | 1.02 | 5.47 | 1.24 | 9.33*** | 0.94 | 5.53 | 1.12 | 8.26*** |
| $D_1^*$ (mm<sup>2</sup>/s) | 0.12 | 1.38 | 0.15 | 3.79*** | 0.032 | 1.80 | 0.042 | 4.65*** |
| $D_2^*$ (mm<sup>2</sup>/s) | | | | | 1.49 | 0.83 | 2.27 | 4.10*** |
| $f_1$ (%) | | | | | 15.89 | 1.50 | 15.80 | 2.26*** |
| $f_2$ (%) | | | | | 13.31 | 1.83*** | 8.35 | 1.18 |
| $f_{1+2}$ (%) | 26.28 | 3.30 | 19.91 | 3.66* | 29.20 | 3.23 | 24.15 | 3.60** |

For the bi-exponential approach, the network took a median of 8.7 minutes to train, with a range of 6.5 to 12.8 minutes, and 40 seconds to inference (1.2×10^−5^ seconds per voxel). For the tri-exponential approach, the network took a median of 12.2 minutes to train, with a range of 3.4 to 16.2 minutes, and 52 seconds to inference (1.5×10^−5^ seconds per voxel). The least-squares fits took substantially longer, with 85 minutes (150×10^−5^ seconds per voxel) for bi-exponential fitting and 166 minutes (300×10^−5^ seconds per voxel) for tri-exponential fitting. Training took a median of 91 epochs before converging.

### Comparison of bi-versus tri-exponential model

The average adjusted R^2^ of the LSQ tri-exponential fit for the ROI_liver_ was 0.990 for the tri-exponential model and 0.984 for the bi-exponential model, with a higher adjusted R^2^ for the tri-exponential model in 33 out of 36 patients. The bi-exponential and tri-exponential R^2^ values of each individual along with example fits of the best and worst performing tri-exponential fit compared to bi-exponential fit can be found in Figure 6.

**Figure 6.**
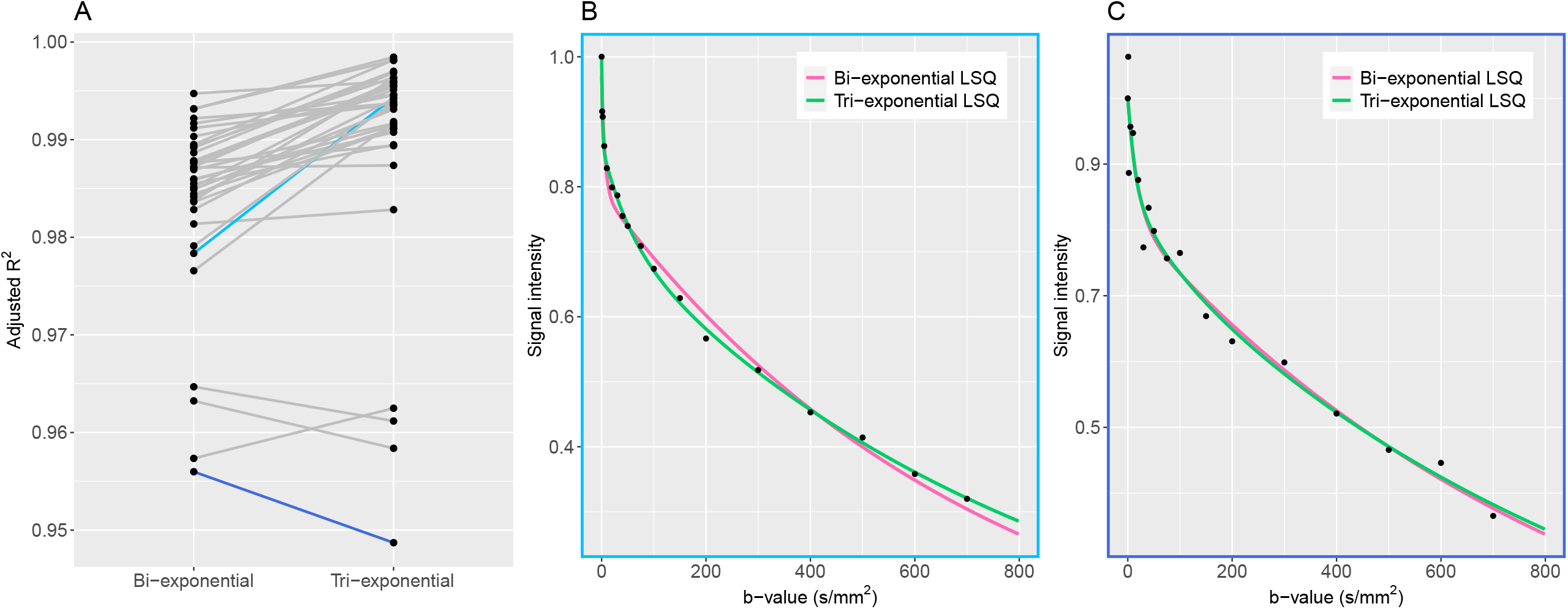
Adjusted R^2^ plots. A: adjusted R^2^ values for the bi- and tri-exponential least-squares models for each individual. The blue lines represent the patients with the largest difference between bi- and tri-exponential fits, with light blue showing a higher R^2^ for the tri-exponential fit and dark blue a higher R^2^ for the bi-exponential fit. B: example plot of the average signal intensity for each b-value and the corresponding bi- and tri-exponential fits from the patient with the largest difference between bi- and tri-exponential fits, favouring the tri-exponential fit. The tri-exponential fit more accurately fits the data points. C: example plot of the average signal intensity for each b-value and the corresponding bi- and tri-exponential fits from the patient with the largest difference between bi- and tri-exponential fits, favouring the bi-exponential fit. Here the data points show more spread, indicating a noisier dataset.

### Correlations with liver histopathology

Figure 7 gives an overview of the Spearman correlations between IVIM parameters and histopathology for all the reported fit methods. Comparing LSQ fits (top 2 rows) with NN fits (bottom two rows), ten correlations are stronger for the NN than LSQ fit, whereas the LSQ finds five that are stronger than the NN. When comparing bi-exponential to tri-exponential models (using bi-exponential *f* and tri-exponential *f*_1+2_), marginal differences between the four models were found. *D* showed higher correlations between histopathology for the bi-exponential model, in particular for the LSQ model. On the other hand, *f* was higher for the tri-exponential LSQ model compared to the bi-exponential model, while the NNs showed overall similar values.

**Figure 7.**
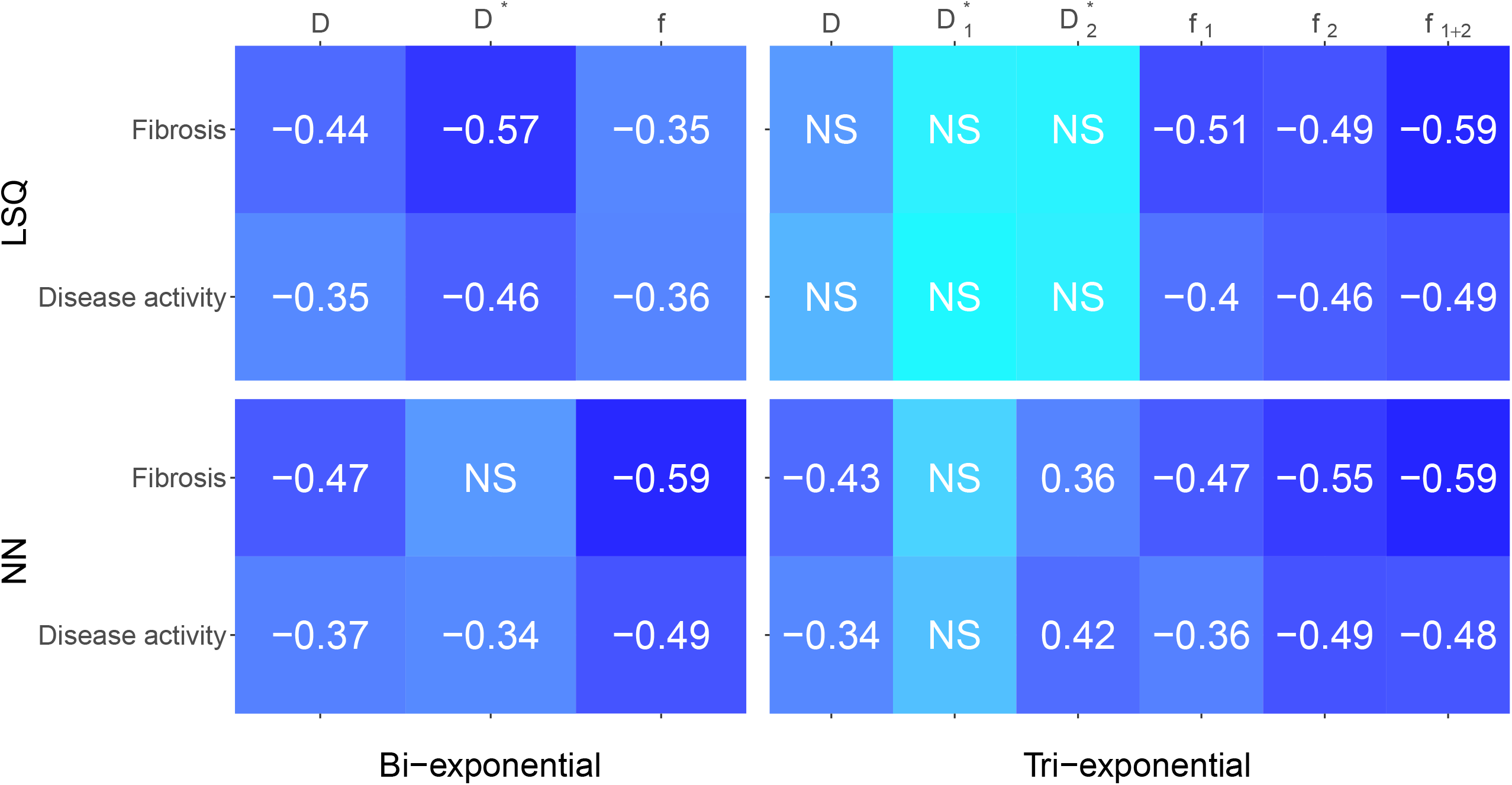
Spearman correlations between histopathological outcomes fibrosis and disease activity versus IVIM parameters for the bi- and tri-exponential least-squares methods (LSQ) and the bi- and tri-exponential neural network methods (NN). Darker blue equates a stronger negative or positive correlation. Only significant values (p < 0.05) are reported, non-significant values are depicted by NS.

#### IVIM_3_-NET versus Liver Fibrosis

IVIM_3_-NET showed a significant decrease in *D* (r_s_ =-0.43, p =0.0097), *f*_1_(r_s_=-0.47, p =0.0036), *f*_2_ (r_s_ =-0.55, p <0.001) and *f*_1+2_ (r_s_ =-0.59, p <0.001) with increasing fibrosis stage. *D*\* was positively correlated with fibrosis stage (r_s_ =0.36, p =0.033). Significant differences in medians between fibrosis stages were seen for *f*_1_ (χ^2^=11.18, p=0.025, df= 4), *f*_2_ (χ^2^=11.00, p=0.027, df= 4) and *f*_1+2_ (χ^2^= 14.43, p= 0.0060), while post-hoc analysis only showed a significant difference between fibrosis stage 0-3 for *f*_1_ and *f*_1+2_ (Figure 8).

**Figure 8.**
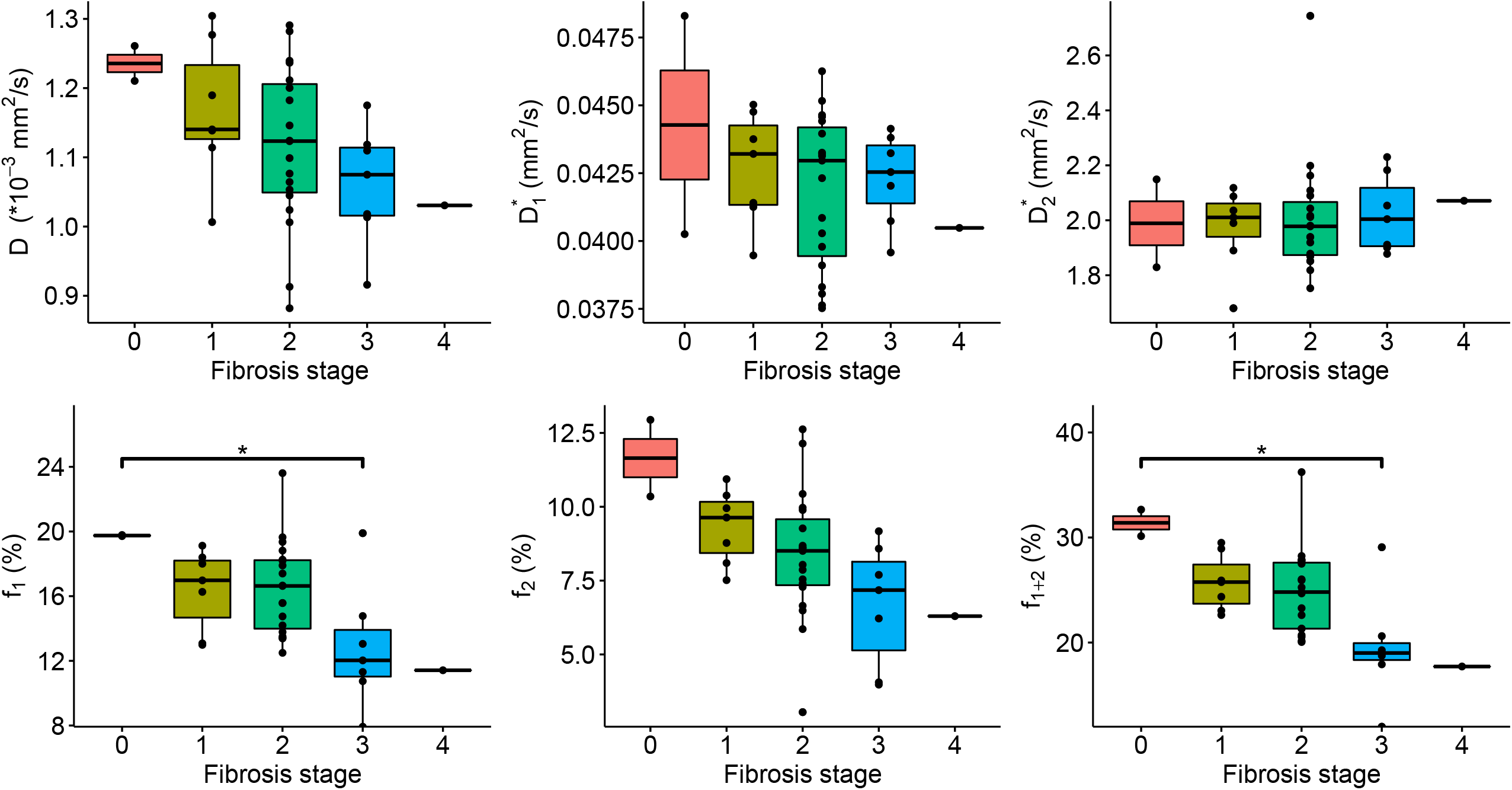
IVIM_3_-NET parameters versus fibrosis grade. All parameters except 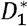; showed a significant correlation with fibrosis, while only *f*_l_ showed a significant difference in medians between fibrosis stage 0 and 3; *f*_l+2_ between stage 0 and 3, as well as stage 1 and 3.

#### IVIM_3_-NET versus NASH Disease Activity

A significant correlation was observed between disease activity grade and all IVIM_3_-NET parameters except 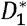 (*D*: r_s_ = -0.35, p =0.040 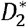: r_s_ = 0.42, p =0.011; *f* : r_s_ = -0.36, p = 0.029; *f*_2_ : r_s_ = -0.49, p =0.0024; *f*_1+2_: r_s_ = -0.48, p = 0.0029) was observed, with *f*_2_ and *f*_1+2_ showing significant differences between disease activity grades (χ^2^= 9.77, p= 0.044, df = 4; χ^2^= 9.97, p= 0.041, df = 4 respectively). Post-hoc analysis, however, did not provide significant differences between individual disease activity grades (Figure 9).

**Figure 9.**
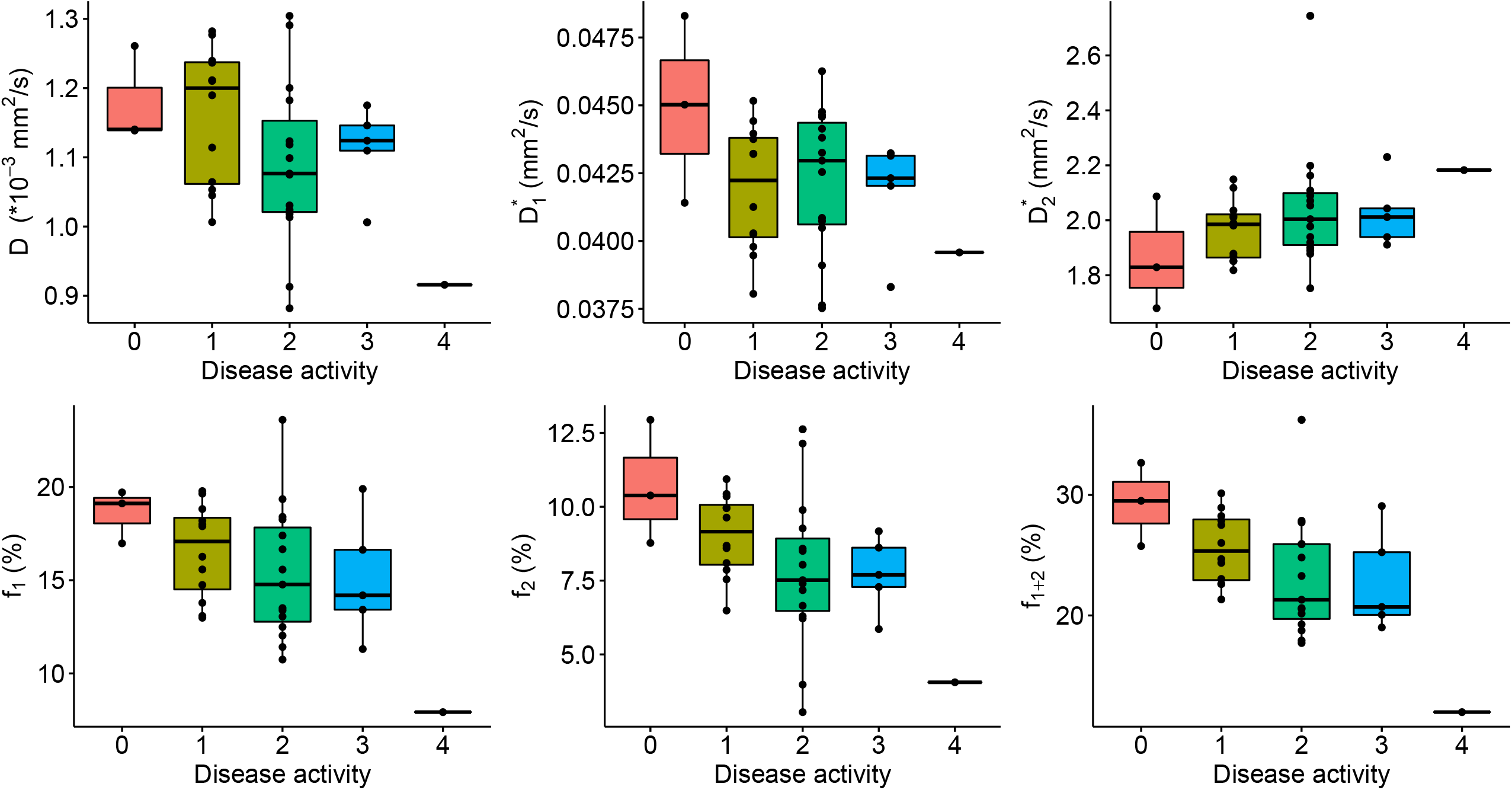
IVIM_3_-NET parameters versus disease activity. All parameters except 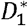 showed a significant correlation with disease activity, yet no significant differences between medians of disease activity grades were found.

#### IVIM_3_-NET versus Hepatic Steatosis

Steatosis grade did not correlate with any IVIM_3_-NET parameter and no significant differences between group medians were found (Supplementary Information Figure S2).

## Discussion

Our IVIM_3_-NET is the first neural network able to provide high-quality tri-exponential IVIM parameter maps and IVIM_3_-NET is shared on our GitHub. We had four main findings: first, the parameter map quality improved with neural network approaches, as both neural networks provided a less noisy fit than the LSQ fits. Second, the neural network was substantially faster and produced parameter maps in times that would allow for immediate on console evaluation. Third, the majority of patients with NAFLD displayed a higher adjusted R^2^ for the tri-exponential model compared to the bi-exponential model, suggesting that the data behaves in a tri-exponential manner. Fourth, correlating IVIM data to liver histology results from patients with varying levels of NAFLD severity showed that both the IVIM_3_-NET and IVIM-NET showed similar diagnostic performance, while generally outperforming both the bi- and tri-exponential least-squares models.

Multiple studies in healthy volunteers support the use of a tri-exponential model for assessing IVIM scans of the liver. For example, Cercueil et al.^14^, Riexinger et al.^25^ and Chevallier et al.^12^ all assessed the use of a tri-exponential model of the liver compared to a bi-exponential model and showed an improvement in the Akaike information criterion for the tri-exponential model amongst others. To our knowledge, we are the first to assess the use of a tri-exponential model with an extra fast diffusion component in patients with liver disease. Our results also support the use of a tri-exponential model, seen by a higher adjusted R^2^ in the majority of subjects. The clinical implications are less clear in our cohort, as both the bi- and tri-exponential models showed comparable correlations with histopathology. However, NAFLD is known to cause vascular changes^26^, thus particularly the evaluation of the additional perfusion parameter *f*_2_ could be of interest, as this signal originates from larger vessels^11^. We (visually) observed a large decrease in *f*_2_ volumes with increasing levels of fibrosis (Figure 4), which could lead to radiological markers for disease severity.

The use of a neural network for the generation of IVIM parameter maps resulted in higher quality images when compared to a least-squares fit, and in a fraction of the time. A higher correlation was found between histopathology and neural network IVIM parameters compared to LSQ fits, highlighting a promising application for the use of the IVIM-NET and IVIM_3_-NET parameters as a non-invasive biomarker in NAFLD. Moreover, with the improved image quality, additional information may be available. Improved image quality allows for the analysis of images in a voxel-wise manner and may be able to provide more insights into disease pathophysiology. The visual decrease of areas with high signal intensity on *f*_1_ and *f*_2_ images with increasing fibrosis grades may, for example, be indicative of the before mentioned vascular changes in NAFLD. Thus voxel-wise assessment of images, such as volume of voxels with higher signal intensities, could be an interesting route to explore. Furthermore, while IVIM studies tend to focus on quantitative measures, improved parameter map quality may allow for qualitative assessment of images, for example aiding diagnosis of liver lesions such as presented in Figure 5.

For healthy volunteers, an overview of average tri-exponential IVIM-values was published in a recent review^11^. Values for *D* ranged between 0.98-1.35 *10^−3^ mm^2^/s which is in line with what we found with the IVIM_3_-NET and higher than our tri-exponential LSQ fit as seen in Table 1. 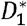 values were similar to values found in our patient cohort, with reported values ranging between 15.4 and 81.3 *10^−3^ mm^2^/s. The largest variation was found in the 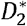 parameter, with values between 270 and 2453 *10^−3^ mm^2^/s. The 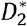 values of both tri-exponential models in our study fit fall in this range, however, the IVIM_3_-NET values were at the high end of the range. The *f*_1_ values of both methods fell within the reported range of 7.8-17.6%, while *f*_2_ was lower than reported values of 10.8-17.1 for the IVIM_3_-NET only.

Discrepancies between values found in this study and previous studies could in part be explained by intrinsic effects caused by NAFLD. We would expect the values in the literature derived from healthy volunteers to be most comparable to patients exhibiting a lesser NAFLD severity, i.e. low levels of fibrosis and disease activity. From Figures 8 and 9 it is clear that 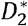 increases with disease activity and fibrosis stage and other parameters including *f*_2_ decrease, suggesting that healthy volunteers could have a lower 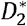 and higher 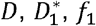 and *f*_2_ than we find in our population.

The wide spread in 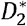 could in part be due to the b-value distribution. Riexinger et al. investigated the effects of the number of b-values acquired, showing that in particular 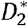 was highly influenced by the number of low b-values due to an increase in fit accuracy, with an increase in 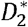 as the number of b-values is increased^13^. The b-value distribution used in this study was optimized for a bi-exponential model and included fewer low b-values than the recommended five to eight b-values < 6 s/mm^2^. Despite this, our IVIM_3_-NET showed 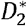 values in the high end of the range of previously published work. This could in part be explained by the fact that the IVIM_3_-NET is relatively uninfluenced by the presence of noise compared to the LSQ fit, however, the effect of b-value distribution will need to be assessed in future work.

Bayesian fitting techniques resulted in comparable parameter map quality to the IVIM-NET for bi-exponential fitting, however, are impractical for tri-exponential fitting due to long calculation times^16^. Setting tighter fit constraints for the least-squares model may result in less noisy parameter maps. However, the same is true for the neural network, which had identical constraints in this work. Our chosen fit constraints were relatively broad to encompass an expected large spread in values between participants, but further work will be required to determine the optimal settings in this population. Finally, denoising the DWI data^27,28^ as a pre-processing step may reduce the noise in the parameter maps of the least-squares fit. However, we opted against denoising as it often introduces blurring of images.

Furthermore, denoising could equally well be performed before the neural networks to further reduce the noise in the output.

In this study, we adapted the IVIM-NET physics-informed deep neural network to include tri-exponential fitting. We chose a physics informed loss function as it allows for training on patient data without inputting any ground-truth answers. Conventional loss functions are also available^17,19^, but they either need to be trained on simulated data, or try to recreate the least-squares fit results in patient data. Therefore, we believe a physics-informed approach has more potential. We tried to minimize the changes to the already optimized IVIM-NET that we adapted the network from, to achieve comparable performance. However, tri-exponential fitting is more challenging and some adjustments were made to address this. The main differences were the implementation of a scheduler for the learning rate and splitting the network in three parts, which we found to improve the tri-exponential fit. Potentially, the network can be further improved in the future by adding spatial awareness with convolutional layers, such as done for e.g. DCE^29^.

In conclusion, the use of a tri-exponential IVIM model can be applied in patients with NAFLD and the IVIM_3_-NET can be used to produce high quality IVIM parameter maps, displaying less noise than a tri-exponential LSQ fit and providing strong correlations with liver histopathology scores, the current gold standard to diagnose and grade NAFLD.

## Data Availability

All data produced in the present study are available upon reasonable request to the authors.

## Abbreviations

NAFLD: non-alcoholic fatty liver disease
IVIM: intravoxel incoherent motion
*D*: diffusion
*D*\*: pseudo-diffusion
*f*: perfusion fraction
*S*0: signal intensity at b=0
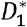: slow pseudo-diffusion
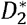: fast pseudo-diffusion
*f*_1_: slow perfusion fraction
*f*_2_: fast perfusion fraction
LSQ: least-squares
ROI: region of interest
IVIM-NET: neural network for bi-exponential IVIM fitting
IVIM_3_-NET: neural network for tri-exponential IVIM fitting
ANCHOR: Amsterdam NAFLD-NASH cohort
SENSE: parallel imaging factor
SPAIR: spectral attenuated inversion recovery
ROI_liver_: region of interest of entire liver excluding large vessels
ROI_SNR_: small region of interest in homogenous liver tissue
SNR: signal-to-noise ratio

## Supplementary Information Figure Legends

**Supplementary Information Figure S1**. Overview of mean and standard error from small homogenous liver tissue region-of-interest (ROI_SNR_) for each individual and each of the four IVIM model parameters.

**Supplementary Information Figure S2**. IVIM_3_-NET parameters versus steatosis. No significant correlations were found between the parameters and steatosis.

