## Supplementary figures and images for "Self-supervised neural network improves tri-exponential intravoxel incoherent motion model fitting in non-alcoholic fatty liver disease"

### Supplementary Information Figure S1

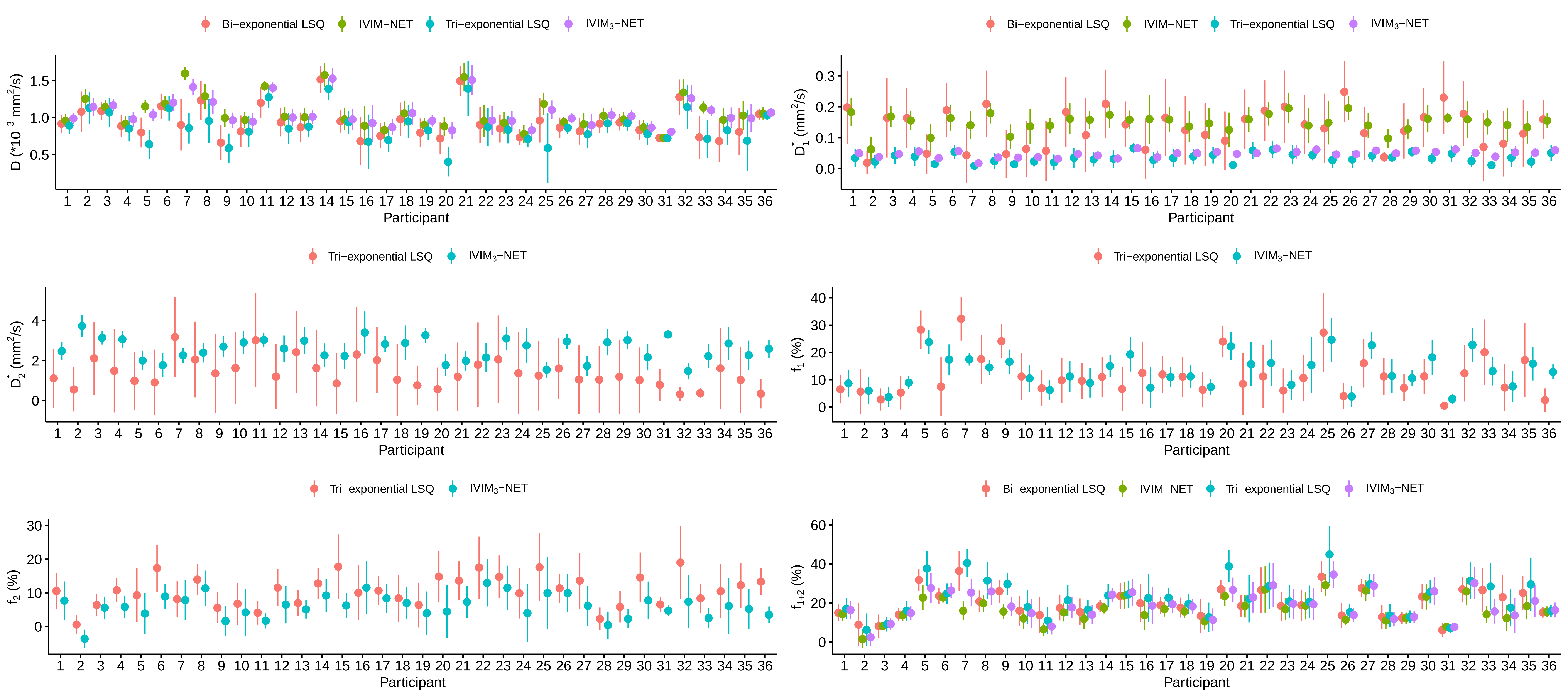

### Supplementary Information Figure S2

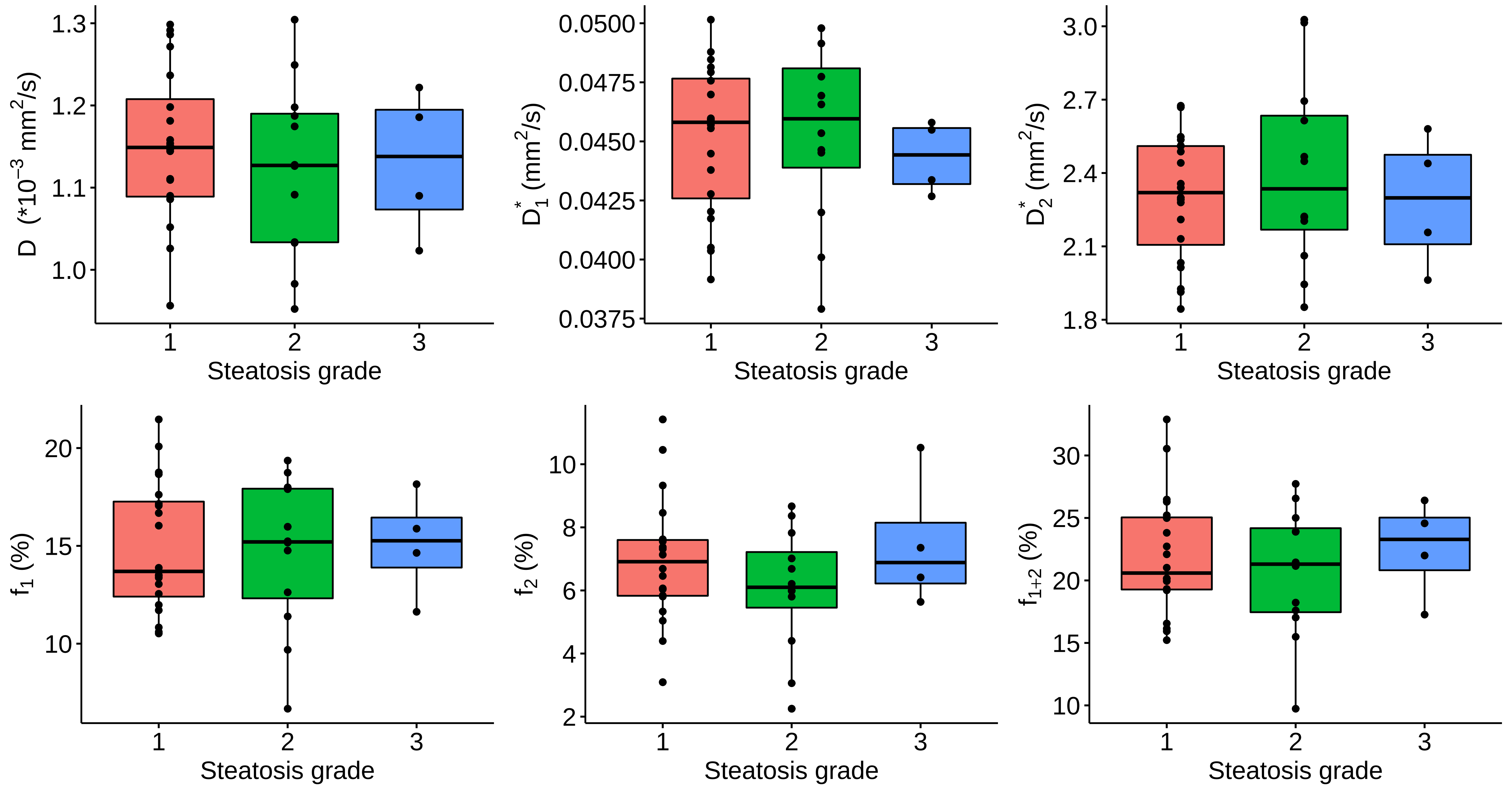
